# Interoperability in NHS Acute Trusts within England: a Situation and Capability Analysis using Freedom of Information requests

**DOI:** 10.1101/2021.10.23.21265418

**Authors:** Sanjay Budhdeo, Chathika K Weerasuriya, Joe Zhang, John P Thomas, Neethu Mariam, Nikhil Sharma, Theodore D Cosco, Oliver T Harrison, Amitava Banerjee

## Abstract

**Background:** Many central initiatives to improve digital maturity and interoperability in the NHS started after 2015. There are few prior assessments of digital maturity and interoperability.

**Methods:** Freedom of Information Act requests were sent to all English Acute NHS Trusts and Clinical Commissioning Groups (CCGs) to obtain information regarding digital maturity according to the Healthcare Information and Management Systems Society (HIMSS) Electronic Medical Record Adoption Mode (EMRAM) scale, and interoperability.

**Results:** One third of Acute NHS Trusts have an EMR that meets requirements for EMRAM stage 6 or above. 17.4% of responding Trusts considered this. 59.1% of responding Trusts stated that their EMR allows for functional interoperability with other (interoperable) EMRs. The majority of responding Trusts had not conferred with other Trusts when making EMR purchasing decisions.

**Discussion:** In order to realise the benefits of digitisation and interoperability, we discuss policy recommendations including actions for local health economies.

## Introduction

Both in the UK and internationally, there is an increasing trend towards the measurement and improvement of the digital maturity of health systems(1–4), with electronic health records (EHR) as the foundation. There is increasing recognition that EHRs are necessary, but insufficient on their own for digital maturity. Interoperability refers to the capability of two or more EHRs to exchange information and make use of that information (5). It is essential for safe and quality care(6), and carries downstream benefits that are vital to implementing the government’s vision for the future of healthcare in the UK(7).

There is widespread recognition of the importance of interoperability. There are three major benefits from efficient data sharing between EHRs and healthcare organisations that would result in improved patient care. First, interoperability allows rapid and reliable access to patient information regardless of speciality, location or system. The Five Year Forward View(8) recognises that multimorbid patients increasingly rely on complex and subspecialised care, taking place across multiple sites at local, regional and national levels. Data-sharing must be improved to facilitate safe and quality care. Second, new approaches in population health, genomics, telehealth and machine learning techniques rely on datasets with high resolution, high fidelity and scale(3,9–11). Interoperability would bring about the linkage of diverse data resources to create large, and well-curated datasets. Third, interoperability of EHR would generate datasets that can catalyse and inform audit processes, identification of variations in care, quality improvement and health policy, at an unprecedented scale. Lack of interoperability therefore limits healthcare professionals, patient care, and health systems.

The call for increasing digitisation in the NHS has increased since 2015. A number of reports have been published recommending change. This includes NHS Five Year Forward View in 2014 (12), the Wachter Review 2016 (13), Life Sciences Industrial Strategy in 2017 (14), Topol Review in 2019 (15), NHS Long Term Plan in 2019 (16). Following the failures of the National Programme for IT (17), and the care.data programme (18), efforts around digitisation and data sharing have moved away from centrally directed and delivered programs, to change within hospital Trusts or local health economies. These initiatives have included Global Digital Exemplars and fast followers (19), Local Health and Care Record Exemplars (20) and the Industrial Challenge Strategy Fund AI Centres (21). Additionally, there have been capability building exercises for provider roles in this area, including the NHS Digital Academy and Topol Fellowships (15).

We had three principle aims in this study. First, we wished to establish the level of digital maturity within the NHS, prior to centrally driven reports and initiatives to encourage digitisation and interoperability in providers. Second, we investigated the extent of data sharing available in the NHS in England through interoperability at this time point. Third, we investigated the extent to which there was coordination of digital resources between Acute Trusts through means other than interoperability, including franchised electronic medical records. Given the challenges of assessing interoperability, we adopted an approach of using Freedom of Information requests to investigate interoperability in NHS Acute Trusts in England in 2015. This enables examination of a base state of interoperability prior to centrally-led policy initiatives to improve digital maturity in the NHS.

## Methods

### Selection of Measures

In order to assess digital maturity we used the Healthcare Information and Management Systems Society (HIMMS) Electronic Medical Record Adoption Model (EMRAM) standard (22), for three reasons. First, this is an international standard that is well recognized and allows for benchmarking of digital maturity in England versus other countries. Second, the standard provides seven clearly demarcated gradations of digital maturity. Third, each gradation has clear criteria to meet, which facilitates self-assessment.

We assessed whether organisations met HIMSS EMRAM stage 6 for two reasons. First, HIMSS divide between Stages 0-5 and Stages 6-7 by including a Stage 6-7 providers list. Second, this dichotomous variable has been previously used in the research literature (23).

### Search Strategy and Data collection

We aimed to collect information from all Acute NHS Trusts and Clinical Commissioning Groups (CCGs) in England. We obtained a list of all Acute NHS Trusts and CCGs from publicly available sources in September 2015: http://www.nhs.uk/servicedirectories/pages/acutetrustlisting.aspx; http://www.england.nhs.uk/south/ccg-trust/; https://www.england.nhs.uk/north/ccg-trust/; https://www.england.nhs.uk/mids-east/ccg-trust/; https://www.england.nhs.uk/london/ccg-trust/; https://www.england.nhs.uk/ccg-details/. We define Acute NHS Trusts as providers of predominantly hospital-based secondary health services. We included trusts that had attained Foundation status, as well as those that had not at the time of sampling. Contact email addresses for officers responsible for Freedom of Information Act (FOIA) requests were obtained from each respective institutional website, or, where unavailable, from a public online repository maintained by mySociety (https://www.mysociety.org). We sent questionnaires (supplementary tables, Table S1 and Table S2) via email to the respective FOIA officers in November 2015. Requests not acknowledged were followed up by repeat email and telephone call. 166 Acute NHS Trusts and 209 CCGs were contacted. The response rate for Acute NHS Trusts was 80%. The response rate for CCGs was 100%.

**Table S1.**
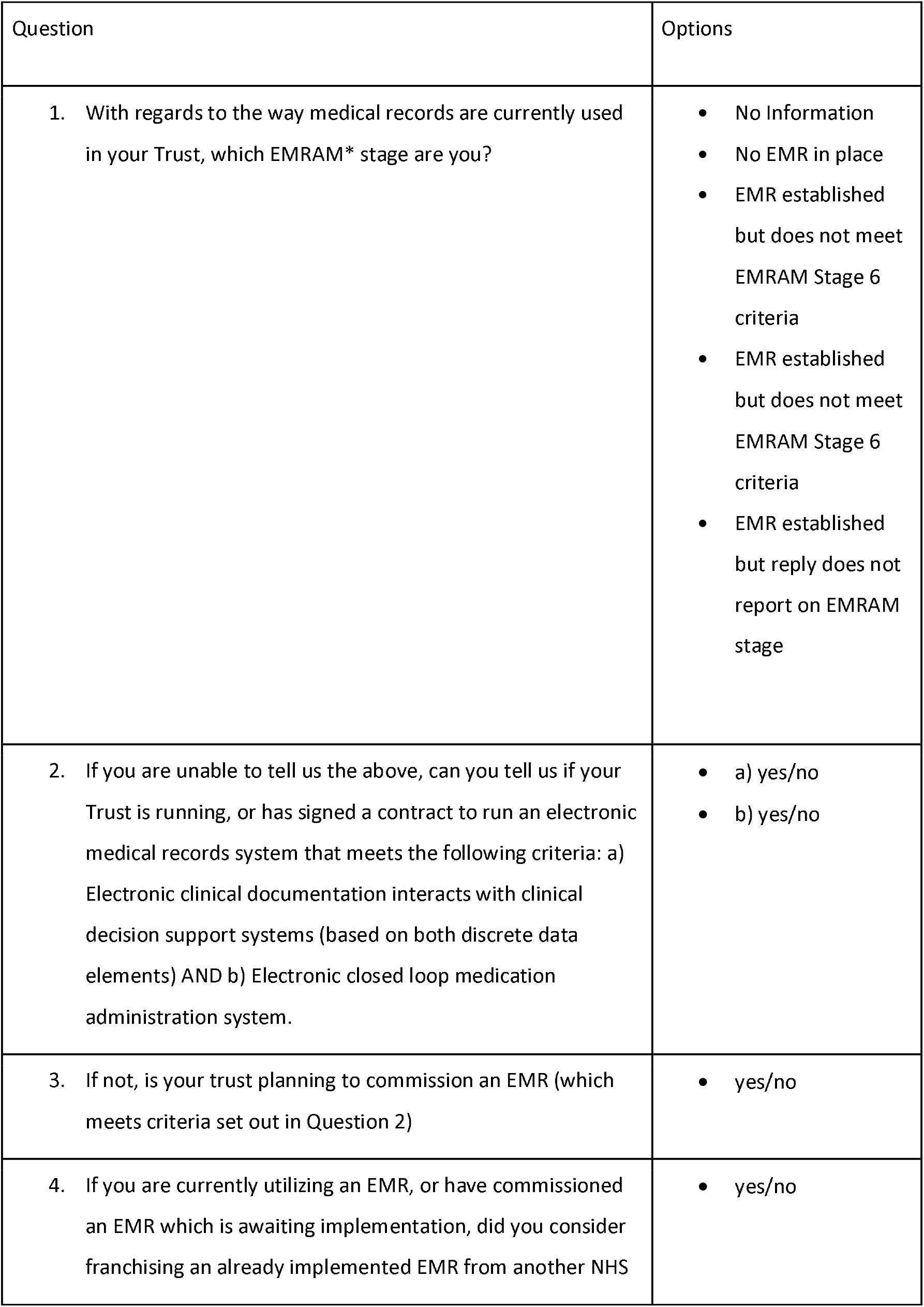
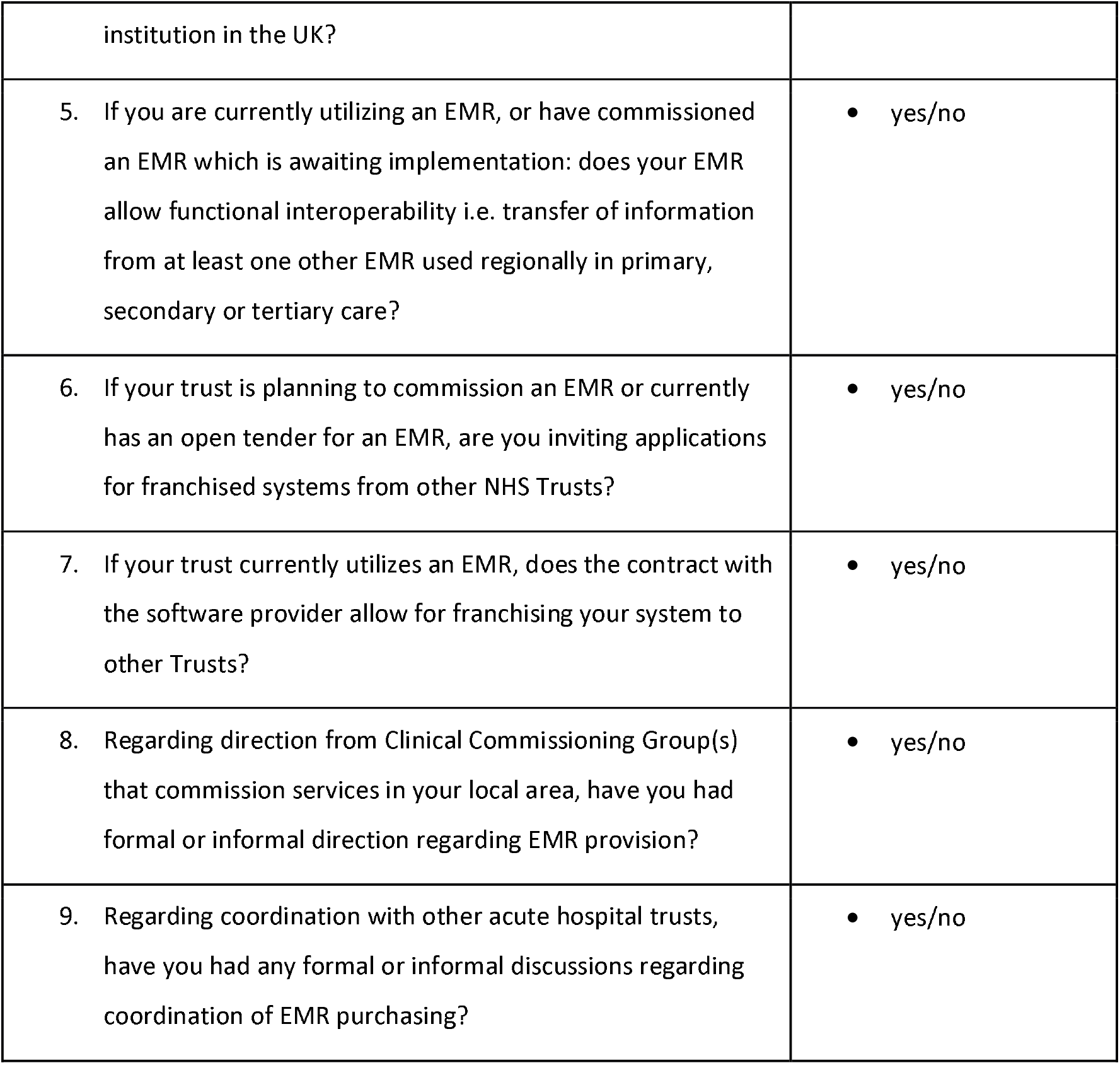
Questionnaire to Acute NHS Trusts

**Table S2.** Questionnaire to Clinical Commissioning Groups

| Question | Options |
| --- | --- |
| 1. Have you had any input, formal coordinating role or informal coordinating role with regards to EMR purchasing and/or use in primary care? | • yes/no |
| 2. Have you had any input, formal coordinating role or informal coordinating role with regards to EMR purchasing and/or use in acute hospital trusts? | • yes/no |
| 3. Do you hold any information about any EMR franchising or | • yes/no |
| coordination arrangements in your geographical area, even if you did not have any input or coordinating role? |  |
| 4. Do you hold records of whether primary care centres in your commissioning area are able to franchise EMR to other primary care centres? | • yes/no |
| 5. Do you hold records of whether acute hospital trusts in your commissioning area are able to franchise EMR to other trusts? | • yes/no |
| 6. Is there functional interoperability (i.e. the ability to transfer of information between EMRs) between two acute trusts within your commissioning area? | • yes/no |
| 7. Is there functional interoperability (i.e. the ability to transfer of information between EMRs) between a primary care organization and an acute hospital trust in your commissioning area? | • yes/no |

Questionnaires were constructed for Acute NHS Trusts (Table S1) and CCGs (Table S2). In this study, EMR capabilities of acute trusts were assessed against the Healthcare Information and Management Systems Society (HIMMS) EMR Adoption Model (24).

### Data Analysis

Data analysis was undertaken in Microsoft Excel 2010 and SPSS version 23 (IBM Corp., 2015).

## Results

Figure 1 shows the EMRAM Stage of Trusts by self-assessment. 39% have an EMR with EMRAM stage less than 6. 33% have an EMR with EMRAM stage 6 or greater. 10% of Trusts have no EMR. 18% provided insufficient information for us to assess status. Table 1 and Table 2 detail the responses from Trusts and CCGs respectively to the questions in Freedom of Information requests summarised in Table S1 and Table S2.

**Table 1:** Responses from Acute NHS Trusts to questions outlined in Table S1. Results from Question 1 and 2 are not included here as they have been used to produce Figure 1.

|  |  | Count | Percentage |
| --- | --- | --- | --- |
| 3. Is your trust planning to commission an EMR? | No | 18 | 13.6% |
|  | Yes | 48 | 36.4% |
|  | NA | 66 | 50.0% |
| 4. If you are currently utilizing an EMR, or have commissioned an EMR which is awaiting implementation, did you consider franchising an already implemented EMR from another NHS institution in the UK? | No | 67 | 50.8% |
|  | Yes | 23 | 17.4% |
|  | NA | 42 | 31.8% |
| 5. If you are currently utilizing an EMR, or have commissioned an EMR which is awaiting implementation: does your EMR allow functional interoperability i.e. transfer of information from at least one other EMR used regionally in primary, secondary or tertiary? | No | 15 | 11.4% |
|  | Yes | 78 | 59.1% |
|  | NA | 39 | 29.5% |
| 6. If your trust is planning to commission an EMR or currently has an open tender for an EMR, are you inviting applications for franchised systems from other NHS trusts? | No | 40 | 30.3% |
|  | Yes | 4 | 3.0% |
|  | NA | 88 | 66.7% |
| 7. If your trust currently utilizes an EMR, does the contract with the software provider allow for franchising your system to other trusts? | No | 54 | 40.9% |
|  | Yes | 21 | 15.9% |
|  | NA | 57 | 43.2% |
| 8. Regarding direction from Clinical Commissioning Group(s) that commission services in your local area, have you had formal or informal direction regarding EMR provision? | No | 60 | 45.5% |
|  | Yes | 62 | 47.0% |
|  | NA | 10 | 7.6% |
| 9. Regarding coordination with other acute hospital trusts, have you had any formal or informal discussions regarding coordination of EMR purchasing? | No | 52 | 39.4% |
|  | Yes | 60 | 45.5% |
|  | NA | 20 | 15.2% |

**Table 2:** responses from CCGs to questions outlined in Table S2

|  |  | Count | Percentage |
| --- | --- | --- | --- |
| 1. Have you had any input, formal coordinating role or informal coordinating role with regards to EMR purchasing and/or use in primary care? | No | 58 | 27.9% |
|  | Yes | 149 | 71.6% |
|  | N/A | 1 | 0.5% |
| 2. Have you had any input, formal coordinating role or informal coordinating role with regards to EMR purchasing and/or use in acute hospital trusts? | No | 137 | 65.9% |
|  | Yes | 68 | 32.7% |
|  | N/A | 3 | 1.4% |
| 3. Do you hold any information about any EMR franchising or coordination arrangements in your geographical area, even if you did not have any input or coordinating role? | No | 167 | 80.7% |
|  | Yes | 38 | 18.4% |
|  | N/A | 2 | 1.0% |
| 4. Do you hold records of whether primary care centres in your commissioning area are able to franchise EMR to other primary care centres? | No | 172 | 82.7% |
|  | Yes | 34 | 16.3% |
|  | N/A | 2 | 1.0% |
| 5. Do you hold records of whether acute hospital trusts in your commissioning area are able to franchise EMR to other trusts? | No | 198 | 95.2% |
|  | Yes | 8 | 3.8% |
|  | N/A | 2 | 1.0% |
| 6. Is there functional interoperability (i.e. the ability to transfer of information between EMRs) between two acute trusts within your commissioning area? | No | 93 | 44.7% |
|  | Yes | 32 | 15.4% |
|  | N/A | 83 | 39.9% |
| 7. Is there functional interoperability (i.e. the ability to transfer of information between EMRs) between a primary care organization and an acute hospital trust in your commissioning area? | No | 50 | 24.0% |
|  | Yes | 129 | 62.0% |
|  | N/A | 29 | 13.9% |

**Figure 1:**
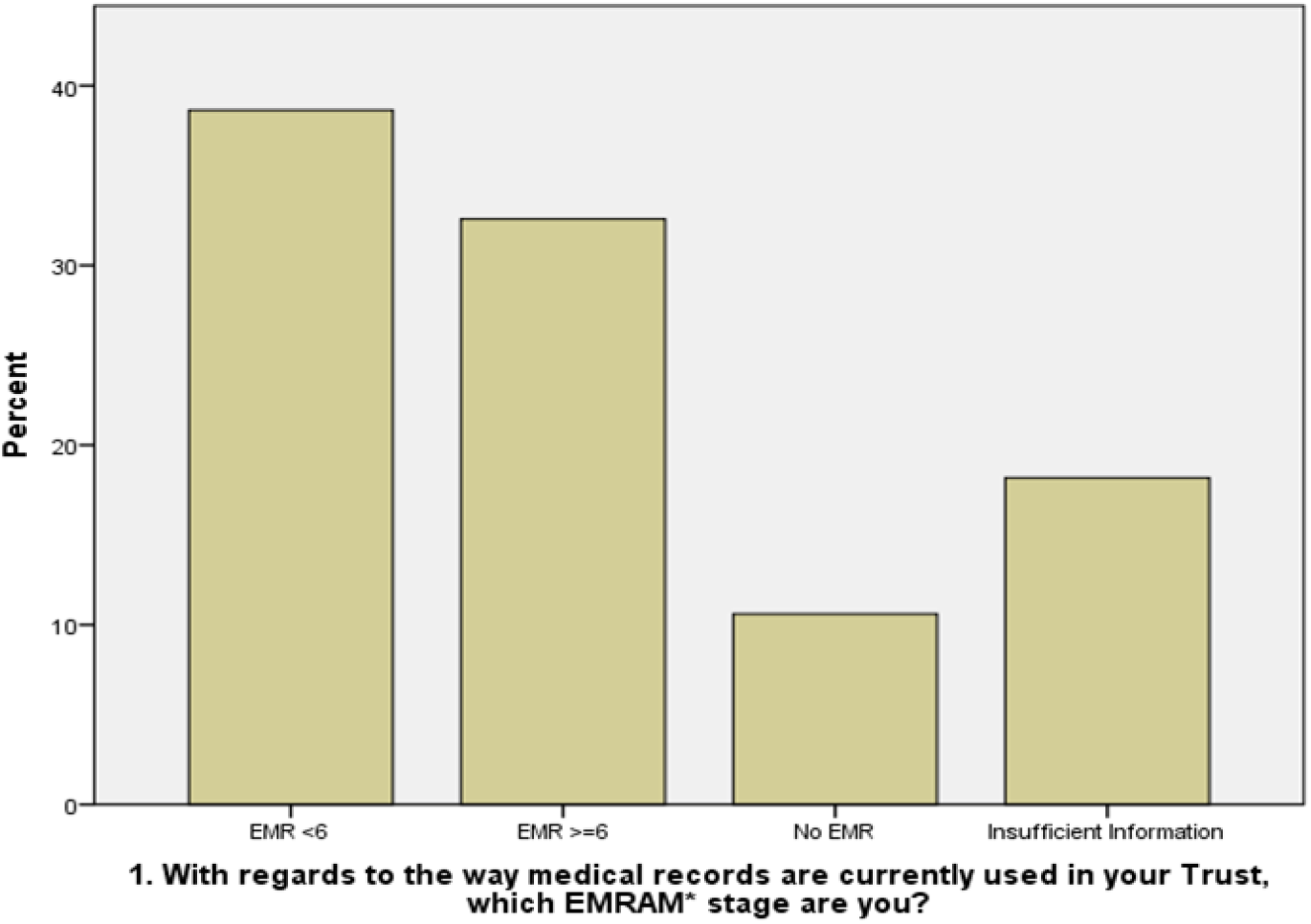
EMRAM Stage of NHS providers (“Trusts”) by self-assessment

## Discussion

### Findings

We have five key findings. First, only one third of NHS Trusts self-reported as having EMR that met requirements for EMRAM stage 6 or above at the time the survey was carried out. Second, franchising of an existing EMR by Trusts was often not considered, only 17.4% of responding Trusts considered this. Franchising is often not considered by payors to aid coordination of care: most CCGs are not aware of the possibility of franchising for primary care (82%) or secondary care (95%) from providers in their local health economy. Third, interoperability between Trusts is poor at present. Only 59.1% of responding Trusts stated that their EMR allows for interoperability with another EMR. Fourth, the majority of responding Trusts had not conferred with other Trusts when making EMR purchasing decisions. Fifth, CCG involvement regarding EMRs is greater in primary care (72% report involvement) versus secondary care (33% report involvement).

### Comparison to research literature

The NHS Digital Maturity Index was compiled in 2015/16 (25). In the NHS digital maturity survey (26), only 56 out of 249 Trusts were able to share digital records with other hospital Trusts, limiting the network benefits across the system. In terms of methodology comparison, institutions were also asked to self-assess their digital maturity in accordance with prescribed standards for this survey. There are two key differences. First, the NHS Digital Maturity Index reviewed acute hospital trusts as well as mental health, community, and ambulance services, providing a broader overview of interoperability. Second, the NHS Digital Maturity Index was designed to review specific policy objectives. In contrast, our study is an independent assessment of NHS EMR use with questions designed to assess internationally validated standards.

KLAS, an independent research organization, has produced a report on vendor level interoperability between electronic medical records (27). This report found only half of surveyed hospital Trusts had EMR systems capable of receiving any outside patient data (28).We acknowledge that provider relationships are only one aspect to interoperability. Vendors have a large influence on ease of interoperability through the development of APIs that permit data sharing between electronic health record systems. In the UK, non-governmental initiatives including the INTEROpen collaboration (29) have moved on this, as well as increasing maturity of FHIR standards, which pertain to transfer data from one data structure to another. Draft standards for FHIR were published in 2014, and updates in 2015, 2017 and 2019.

Analysis of Hospital Episode Statistics which mapped networks of hospitals which shared patients has confirmed that interoperability within local health economies would solve most patient data sharing problem, without recourse to national coordination (30). There is currently limited alignment of EMR systems locally, strengthening our recommendations regarding franchising (31).

### Explanation of findings and implications

Our findings suggest difficulty in uptake of EMRs and in interoperability when measured in 2015, prior to central government drive through various reports and initiatives introduced since 2015. It is unclear what causal factors have meant that central drive has improved digital maturity. It may be the ability to prioritise provider agendas, funding, or culture change. It is most likely that causes are multifactorial and interdependent. Most CCGs did not deliver on coordinating electronic health record provision within local health economies. There were some successful local services have been able to facilitate functional interoperability, for example in East London (32) (33) and West London (34). Central processes have improved digital maturity. Initiatives to encourage interoperability such as LHCREs have been more recent, so their success cannot yet be judged. As another NHS reorganisation is imminent, and there is an increasing role for the Department of Health, consideration of the lessons from digital maturity and interoperability may be useful.

In the United States, the development of HIEs this has been encouraged by federal funding as part of the The Health Information Technology for Economic and Clinical Health (HITECH) Act (35), reinforcing the utility of a central drive for change backed my funding, with local implementation. The care.data programme shows problems of initiatives set up and run from the centre (18). Perhaps locally run projects with incentivization and oversight from the Department of Health will work well in the future.

Without far greater interoperability, it is more challenging and resource-intensive to build joined-up data assets for research. This in turn creates challenges for learning and deployment artificial intelligence tools in healthcare, which is a particular focus of the current government. The learning health system model cannot be achieved without better interoperability (36).

#### Study limitations

The key limitations of this study pertain to time course: data were collected in November 2015; the current status of EHR implementation has advanced beyond our findings (37). However, our aim was to establish an understanding of interoperability prior to newer centrally driven policy changes, and our study produces a foundation from which to assess how recent policies may have promoted the development of interoperability in the intervening time.

The EMRAM 6 digital maturity measure, and interoperability measures, rely on Trust self-report via Freedom of Information requesting. Responders were not presumed to be technical experts in this field, and our results rely on their seeking relevant expertise within Trusts where necessary. Our binary assessment of ‘interoperability’ – data exchange between two organisations for one system – was purposefully simplified to mitigate for potential lack of technical expertise.

### Further work

The conclusions from this analysis could be tested using a longitudinal analysis of interoperability change over time. This could be taken further by cluster analysis within local health economies where interoperability between different players matters the most in terms of maximisation and efficient delivery of patient outcomes.

In addition to research in the NHS context, research on the drivers for interoperability in health systems internationally should be continued. A variety of methods could be used, quantitative and qualitative, to try and understand this.

## Data Availability

All data produced via Freedom of Information Act requests are available online at https://www.whatdotheyknow.com

## Acknowledgements

The authors are grateful for the work of Archita Srinivasan and Matthew Hague in data collection

## Notes

### Competing Interest Statement

SB owns stock in Owkin Inc.

### Funding Statement

This study did not receive any funding

